# Arsenic exposure is associated with elevated sweat chloride concentration and airflow obstruction among adults in Bangladesh: a cross sectional study

**DOI:** 10.1101/2024.09.25.24314390

**Authors:** Mi-Sun S. Lee, Crystal M. North, Irada Choudhuri, Subrata K. Biswas, Abby F. Fleisch, Afifah Farooque, Diane Bao, Sakila Afroz, Sadia Mow, Nazmul Husain, Fuadul Islam, Md Golam Mostafa, Partha Pratim Biswas, David S. Ludwig, Subba R. Digumarthy, Christopher Hug, Quazi Quamruzzaman, David C. Christiani, Maitreyi Mazumdar

**Affiliations:** Department of Environmental Health, Harvard T.H. Chan School of Public Health, Boston, MA USA; Division of Pulmonary and Critical Care Medicine, Massachusetts General Hospital, Boston, MA, USA; Department of Medicine, University of Pittsburgh Medical Center, Pittsburgh, PA, USA; Department of Molecular and Cell Biology, University of Connecticut, Storrs, CT, USA; Center for Interdisciplinary Population Health Research, MaineHealth, Portland, ME USA; Pediatric Endocrinology and Diabetes, Maine Medical Center, Portland, ME USA; Department of Neurology, Boston Children’s Hospital, Boston, MA USA; Dhaka Community Hospital Trust, Dhaka Bangladesh; Department of Biochemistry, Bangabandhu Sheikh Mujib Medical University, Dhaka, Bangladesh; New Balance Obesity Prevention Center, Boston Children’s Hospital, Boston, MA USA; Thoracic Imaging and Intervention Division, Massachusetts General Hospital, Boston, MA USA; Consultant, Brookline, MA, USA

## Abstract

Arsenic is associated with lung disease and experimental models suggest that arsenic-induced degradation of the chloride channel CFTR (cystic fibrosis transmembrane conductance regulator) is a mechanism of arsenic toxicity. We examined associations between arsenic exposure, sweat chloride concentration (measure of CFTR function), and pulmonary function among 285 adults in Bangladesh. Participants with sweat chloride ≥ 60 mmol/L had higher arsenic exposures than those with sweat chloride < 60 mmol/L (water: median 77.5 µg/L versus 34.0 µg/L, *p* = 0.025; toenails: median 4.8 µg/g versus 3.7 µg/g, *p* = 0.024). In linear regression models, a one-unit µg/g increment in toenail arsenic was associated with a 0.59 mmol/L higher sweat chloride concentration, p < 0.001. We found that toenail arsenic concentration was associated with increased odds of airway obstruction (OR: 1.97, 95%: 1.06, 3.67, *p* = 0.03); however, sweat chloride concentration did not mediate this association. Our findings suggest that sweat chloride concentration may be a novel biomarker for arsenic exposure and also that arsenic likely acts on the lung through mechanisms other than CFTR dysfunction.

## INTRODUCTION

Up to 220 million people in as many as 70 countries are exposed to elevated arsenic levels through contaminated groundwater.^1,2^ Arsenic exposure has been associated with lung dysfunction,^3–8^ respiratory symptoms,^9–11^ bronchiectasis,^12^ and increased tuberculosis-related^13^ and respiratory-related mortality.^14^ One proposed mechanism for arsenic’s effects on the lung is impairment of mucociliary clearance.

The cystic fibrosis transmembrane conductance regulator (CFTR) is a cyclic adenosine monophosphate-regulated chloride channel in the apical membrane of airway epithelial cells. CFTR plays an essential role in mucociliary clearance by establishing an osmotic gradient across the airway epithelium that promotes fluid secretion.^15–17^ Mutations in *CFTR*, the gene that encodes CFTR, cause cystic fibrosis, a rare autosomal recessive disease that affects the lungs and digestive system. Cystic fibrosis is also associated with a diminished immune response to some gram-negative bacteria; a similarly-diminished immune response is induced by arsenic exposure.^18,19^

Recent studies in killfish demonstrate that arsenic induces CFTR ubiquitinylation and degradation.^20,21^ In human airway epithelial cell culture, arsenic induces an increase in multiubiquitinlyated CFTR, which results in CFTR degradation and reduced CFTR-mediated chloride secretion.^22^ Together, these reports demonstrate that arsenic impairs CFTR function and support the hypothesis that there is a shared pathobiologic mechanism driving both cystic fibrosis and arsenic-induced lung disease.

In humans, CFTR function is directly measured by a sweat test, which is the gold standard diagnostic test for cystic fibrosis.^23^ CFTR mediates chloride resorption from sweat; in the absence or impaired function of CFTR, sweat chloride concentration is elevated. In a recent study of 100 adults in Bangladesh – a country with among the highest arsenic exposure in the world – water and toenail arsenic concentrations were higher among adults with increased sweat conductivity (a marker for elevated sweat chloride concentrations), and those with the highest sweat conductivity did not have *CFTR* mutations consistent with a genetic diagnosis of cystic fibrosis.^24^ This finding in humans supports the hypothesis that arsenic induces CFTR degradation, as was seen in experimental models, and that CFTR degradation may be a mechanism of arsenic toxicity.

Although sweat conductivity is correlated with sweat chloride concentration, the latter remains the gold standard for identifying CFTR dysfunction.^25^ Therefore, to evaluate the relationship between arsenic exposure, CFTR dysfunction, and lung function, we measured arsenic levels, sweat chloride concentrations, and lung function among adults in Bangladesh. Our study aimed to test the hypotheses that higher arsenic exposure is associated with CFTR dysfunction as assessed by sweat chloride concentrations, and that CFTR dysfunction mediates the relationship between arsenic exposure and lung dysfunction.

All protocols were reviewed and approved by the institutional review boards (IRBs) of Boston Children’s Hospital (BCH) (Protocol number: IRB-P00024501), the Dhaka Community Hospital Trust (DCH), and the Bangabandhu Sheikh Mujib Medical University (BSMMU) (Protocol number: BSMMU/2019/3739). The Harvard T.H. Chan School of Public Health (HSPH) and Massachusetts General Hospital (MGH) relied on BCH IRB review. All participants provided written informed consent.

## METHODS

### Study participants

From May 2018 through November 2022, we enrolled 300 adults from a total of 1800 adults who had participated in studies of arsenic and skin lesions between 2001 and 2003 in Pabna, Bangladesh.^26^ We began by creating a list of potential participants using random digit assignment and contacted each potential participant in order on the list until we had reached our target enrollment. All participants from the 2001-2003 study were initially considered eligible for recruitment. Exclusion criteria included evidence of lung cancer or active pulmonary tuberculosis on chest radiography.

### Arsenic exposure measurements

We collected drinking water samples from each participant’s home. Water arsenic concentrations were measured in the Environmental Engineering Laboratory at the Bangladesh University of Engineering and Technology using graphite furnace atomic absorption spectrometry (GF-AAS).^27^ Drinking water arsenic concentrations from the 2001-2003 study were measured using inductively coupled plasma mass spectrometry (ICP-MS) by the Environmental Laboratory Services (North Syracuse, NY, USA).^26^

We collected toenail clippings from all ten toes on a white sheet of paper, placed them in a small coin envelope (ULINE Model No. S-7798) and stored them at room temperature. The Dartmouth Trace Element core facility measured arsenic in toenails using ICP-MS.^28^ Arsenic concentrations in toenail clippings from the 2001-2003 study were measured at the Harvard T.H. Chan School of Public Health using similar ICP-MS protocols.^29^

### Sweat chloride measurements

We performed sweat collection and sweat chloride testing according to established guidelines.^30^ Briefly, we obtained sweat from the ventral surface of both forearms of each participant. After cleaning the skin with distilled water, we stimulated sweat secretion by iontophoresis (total applied current 1.5 mA, 50µA/cm^2^) for five minutes using 0.5% pilocarpine gel disks (Pilogels^®^) and sweat was collected for 30 minutes in a Macroduct^®^ Sweat Collection System (Wescor Biomedical Systems, Logan, UT). The laboratory measured sweat chloride concentrations within 48 hours at the Department of Biochemistry, Bangabandhu Sheikh Mujib Medical University (BSMMU) using a Dimension^®^ RxL Max^®^ Clinical Chemistry Analyzer (Siemens Healthcare Diagnostics Inc., Newark, DE) with QuikLYTE^®^ Integrated Multisensor Technology (IMT). The LOD and coefficient of variation (CV) were 10 mmol/L and 1.1%, respectively. Samples with sweat volumes less than 15µL were considered insufficient for analysis.

### DNA Isolation and *CFTR* Sequence Analysis

For participants with sweat chloride concentrations ≥ 60 mmol/L, we retrieved archived DNA samples that were obtained in the 2001-2003 study. DNA had been extracted from whole blood samples using the Puregene DNA Isolation kit (Gentra Systems, Minneapolis, MN) and then stored at −80°C. We sent retrieved DNA samples to Ambry Genetics^®^ (Ambry Test: CF; Aliso Viejo, CA) for *CFTR* sequence analysis and deletion/duplication analysis.^31^

### Pulmonary Function Testing (PFT) and questionnaires

We measured lung function using EasyOne® Air handheld spirometers (Medical Technologies, Andover, MA). All tested participants had abstained from smoking for at least 45 minutes prior to testing and completed testing in the seated position after loosening any tight-fitting clothing. Measured parameters included forced expiratory volume in one second (FEV_1_) and forced vital capacity (FVC), and all participants repeated PFTs after receiving four puffs of salbutamol (Beximco Pharmaceuticals, Bangladesh) and waiting for 10 minutes. Daily quality control procedures included multi-flow calibration confirmation using a 3-litre syringe (ndd Medical Technologies, Andover, MA). We stopped performing PFTs in March 2020, midway through participant enrollment, for the safety of participants and study staff because of the COVID-19 pandemic.

Two study investigators manually reviewed all PFTs for ATS acceptability and reproducibility standards.^32,33^ We included PFTs in the analytic cohort if there were at least two acceptable maneuvers and both the FEV_1_ and FVC from the two best trials were within 200mL of one another. We defined airflow obstruction as post-bronchodilator FEV_1_/FVC < 0.7.

We used structured questionnaires to collect information on age, education, occupation, smoking history, and medications. We characterized respiratory symptoms using the American Thoracic Society’s Division of Lung Disease questionnaire (ATS-DLD 1978).^34^ We measured height and weight at the study visit.

### Chest Radiographs

All participants underwent posteroanterior and lateral chest radiographs performed by a trained radiology technician at DCH. All radiographs were evaluated by a DCH staff radiologist who was unaware of arsenic exposure, sweat chloride, or PFT results. The staff radiologist evaluated the radiographs for the presence of parenchymal, bony, or cardiac abnormalities. For participants who were diagnosed with airflow obstruction, a senior radiologist provided additional imaging review using a structured data collection form.

### Statistical Analysis

We first examined all variables using descriptive statistics and assessed potential differences in characteristics between groups using *t*-tests for continuous variables and Chi-square tests for categorical variables. We computed Spearman correlation coefficients (*r*) between water and toenail arsenic concentrations. For samples below the limit of detection (LOD), we estimated water arsenic concentrations as LOD/2. We used the average of the toenail arsenic concentrations from the 2001-2003 and 2018-2022 visits to estimate long-term arsenic exposure. Our primary exposure of interest was toenail arsenic concentration. We modeled toenail arsenic using untransformed values and interquartile ranges (IQR).

We used multivariable linear regression to characterize the relationship between arsenic and sweat chloride. We evaluated for potential confounding effects by age, sex, smoking status (current, former, and never), and education (able to write, primary education, middle school or above) by placing each variable in the models individually and assessing for change in the regression coefficient for the arsenic measure.

We also used logistic regression to estimate the relationship between toenail arsenic concentration and the odds of having a sweat chloride concentration ≥ 60 mmol/L. We chose this threshold to be consistent with diagnostic criteria for cystic fibrosis. We considered toenail arsenic concentration our primary measure of arsenic exposure. In sensitivity analyses, we tested the above-described relationships using lower sweat chloride thresholds (≥ 50 mmol/L, ≥ 40 mmol/L, and ≥ 30 mmol/L), as these lower thresholds are sometimes used to identify those who need additional evaluation for cystic fibrosis.^11^

We then characterized the relationship between toenail arsenic and lung function using multivariable linear regression (FEV_1_, FVC, FEV_1_/FVC) or logistic regression (FEV_1_/FVC < 0.7), adjusting for age, sex, smoking status, education, and height (in centimeters). Lastly, we employed mediation analyses to estimate the degree to which sweat chloride mediates the relationship between long-term arsenic exposure and lung function, adjusting for age, sex, smoking status, education, and height.^35^ Results are presented as regression coefficients (b with 95% confidence interval [95% CI]) for continuous outcomes and odds ratio (OR with 95% CI) for dichotomous outcomes per IQR increment in toenail arsenic concentrations. In sensitivity analyses, we restricted analyses to never smokers to rule out the potential masking effects of smoking on the relationship between arsenic and lung function. We performed statistical analyses using SAS (version 9.4; SAS Institute, Inc.), R (version 4.2.2; R Development Core Team), and RStudio (version 2022.12.0+353) software.

## RESULTS

### Study population

Of the 300 enrolled participants, 31 participants were excluded due to missing toenail clippings (n=10) or insufficient sweat volume (n=21), for a total of 269 (90%) participants. Participant characteristics are shown in **Table 1**. Of these 269 participants, 166 (62%) completed PFTs that met ATS acceptability and reproducibility criteria prior to the COVID19-related shutdown. The 166 participants who underwent PFTs were more frequently women (53% vs 33.6% p<0.001) and generally less educated (p=0.02) than the 134 participants for whom PFTs were not performed; otherwise there were no significant differences between these groups (**Supplementary Table 1**).

### Arsenic concentrations

The distributions of drinking water and toenail arsenic concentrations are shown in **Table 2**. Overall, drinking water and toenail arsenic concentrations in 2018-2022 were lower than those observed in 2001-2003 (water median 41.0 µg/L in 2018-2021 compared with 71.2 µg/L in 2001-2003), reflecting the impact of arsenic awareness and reduction activities. We observed no significant sex differences in drinking water or toenail arsenic concentrations. All toenail arsenic concentrations were above the LOD of 0.002 µg/g. Toenail arsenic concentrations were moderately correlated with drinking water arsenic concentrations (*r* = 0.66, *p* < 0.0001).

### Sweat chloride concentrations

Mean ± SD sweat chloride concentration was 49.1 ± 16.8 (median: 48.0 mmol/L, range: 18.0 to 98.0 mmol/L). A total of 69 (25.7%) participants had sweat chloride concentration ≥ 60 mmol/L. Sweat chloride concentrations were higher in men than women (51.6 ± 16.8 vs. 46.1 ± 16.4, *p*=0.007) and higher among smokers vs. never smokers (52.6 ± 16.5 vs. 47.6 ± 16.8). There were no significant associations between sweat chloride concentration, participant’s age and education. (**Supplementary Table 2)**.

### *CFTR* Sequence Analysis

Of the 69 participants with sweat chloride ≥ 60 mmol/L, 45 (65.2%) underwent *CFTR* sequence analysis. None of the DNA samples tested revealed homozygous pathogenic mutations in *CFTR*. Six participants had heterozygous findings consistent with classification as cystic fibrosis carriers (**Supplementary Table 3**). These findings suggest that the elevated sweat chloride concentrations we observed were not caused by *CFTR* mutations that lead to absent or abnormal CFTR protein production.

### Pulmonary function tests, respiratory symptoms, and chest imaging

No participant was found to have lung cancer, active pulmonary tuberculosis, cardiac or bony abnormalities on chest films. Of the 166 who underwent PFTs, mean ± SD was 1.97 ± 0.54 L for FEV_1_, 2.57 ± 0.67 for FVC, and 0.77 ± 0.09 for FEV_1_/FVC (**Table 3**). Airflow obstruction was present among 19.9% (n=33), most of whom were men (27 of 33, 82%). Most of the 27 men with airflow obstruction (n=23, 85%) had a history of smoking, while none of the six women with airflow obstruction reported a history of smoking. A total of 55 participants (33.1%) endorsed respiratory symptoms, which included wheezing, shortness of breath or coughing. Respiratory symptoms were more prevalent among those with airflow obstruction; 57.6% of people with airflow obstruction reported respiratory symptoms compared to 27.1% of people without airflow obstruction. Among the 33 participants with airflow obstruction identified on PFTs, additional review of chest imaging showed evidence of bronchiectasis in 5 individuals.

### Arsenic and sweat chloride

To evaluate sweat chloride’s potential use as a biomarker, we present the most parsimonious models including modeling arsenic on a natural scale for ease of interpretation. Concurrent drinking water and toenail arsenic concentrations were higher among participants with sweat chloride ≥ 60 mmol/L compared to < 60 mmol/L (water: median 77.0 µg/L versus 34.0 µg/L, *p* = 0.025; toenails: median 4.8 µg/g versus 3.7 µg/g, *p* = 0.024) (**Figure 1**). A similar pattern was seen for long-term arsenic concentration (**Supplemental Figure 1).** In models adjusted for sex and smoking status, a one-unit µg/g increment in toenail arsenic concentration was associated with a 0.59 mmol/L higher sweat chloride concentration, p< 0.001, and a one-unit µg/g increment in toenail arsenic concentration was associated with greater odds of having a sweat chloride concentration ≥ 60 mmol/L (OR: 1.06, 95% CI, 1.01, 1.11). We found similar findings at lower thresholds of sweat chloride. Models examining varying sweat chloride cutoffs are shown in **Supplemental Figure 2** and **Supplemental Figure 3**. In models fully adjusted for covariates, an IQR (6.6 µg/g) increment in long-term exposure to toenail arsenic was associated with a 3.2 mmol/L (95% CI: 0.57, 5.85, *p =* 0.017) higher sweat chloride concentration (**Supplementary** Figure 4).

**Figure 1:**
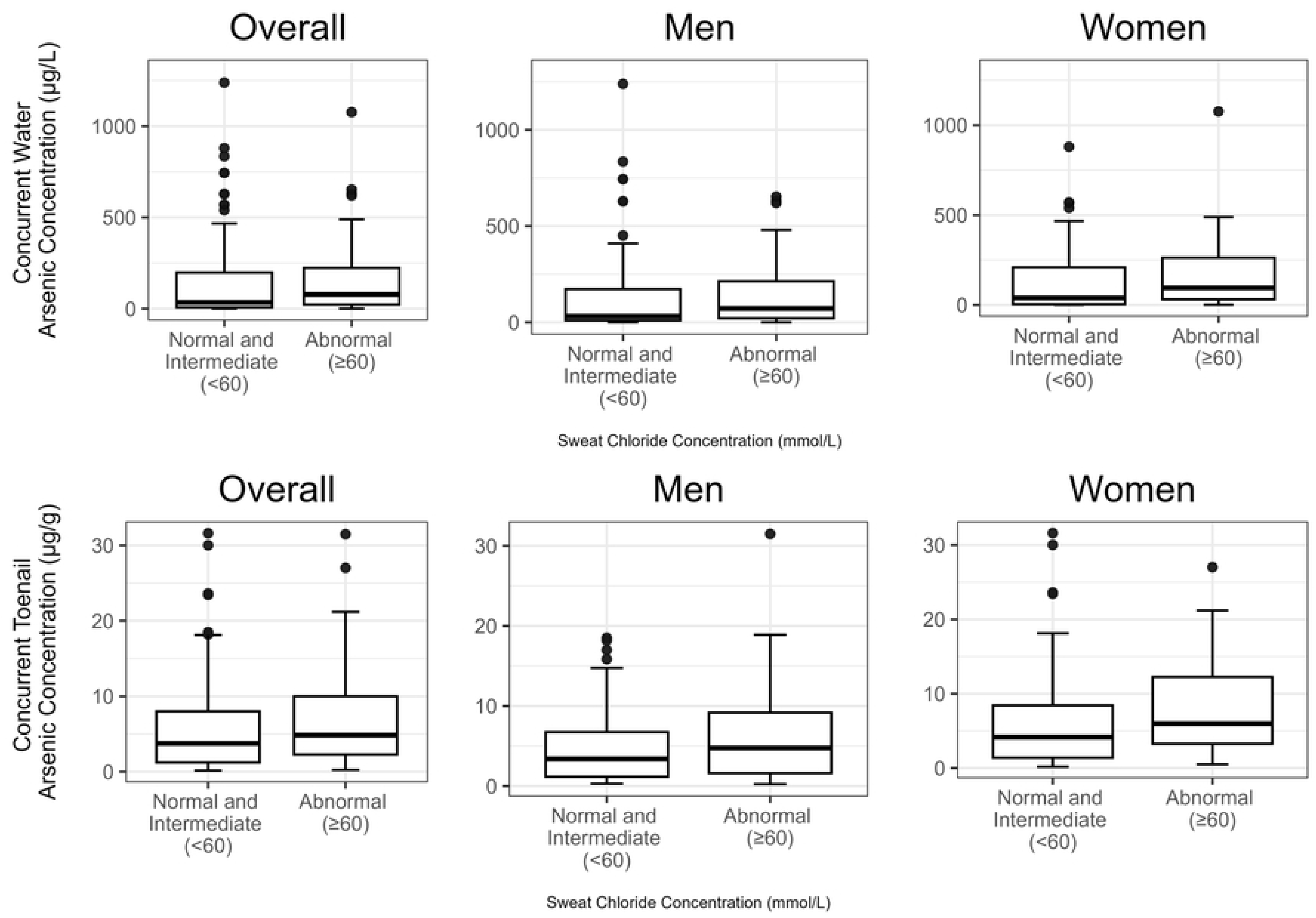

### Arsenic and pulmonary function tests

In adjusted models, an IQR increment in toenail arsenic concentration at each timepoint was associated with nonsignificant reductions in FEV_1_, FVC, and FEV_1_/FVC (**Table 4**). An IQR increment in toenail arsenic concentration from 2001-2003 was associated with greater odds of airflow obstruction (toenail arsenic 2001-2003 OR: 1.57, 95% CI: 1.03, 2.40, *p* = 0.03; long-term toenail arsenic (OR: 1.97, 95%: 1.06, 3.67, *p* = 0.03). Results were similar when we restricted the analytic cohort to never smokers (**Supplementary Table 4**).

### Mediation analysis

Mediation analyses indicate that there is no evidence of sweat chloride as a mediator of the relationship between toenail arsenic concentrations and airflow obstruction (**Supplementary** Figure 4).

## DISCUSSION

Our study demonstrates that 1) elevated sweat chloride concentration, the diagnostic hallmark of cystic fibrosis, is found among individuals exposed to arsenic through drinking water, 2) individuals with sweat chloride concentrations ≥ 60 mmol/L did not have a genetic diagnosis of cystic fibrosis, 3) long-term arsenic exposure as estimated by toenail arsenic concentration is associated with airflow obstruction, and 4) sweat chloride concentration did not mediate the association between arsenic and airflow obstruction. These findings suggest that high sweat chloride may be a novel biomarker of arsenic exposure, but that CFTR dysfunction is unlikely to be the primary mechanism of arsenic-associated lung disease.

Our study has multiple implications for screening of arsenic-related disease in countries such as Bangladesh. Sweat tests may be an effective, inexpensive, point-of-care screening test for arsenic exposure. As we have demonstrated here and has been shown elsewhere,^24^ it is possible to perform sweat tests in rural areas without expensive laboratory equipment. Another implication of our finding is that diagnoses of cystic fibrosis made only by sweat tests (i.e., without genetic testing, as is done in many low and middle income countries including Bangladesh^36^) may be false positives – that is, individuals’ elevated sweat chloride concentrations may be a result of high arsenic exposure and not genetic disease.

Our findings showing associations between arsenic and airway obstruction are consistent with the published literature describing the relationship between arsenic exposure and lung dysfunction. Cross-sectional and cohort studies among childhood and adult populations in China, Mexico, and the United States consistently demonstrate relationships between arsenic exposure and restrictive patterns on lung function testing that are suggestive of restrictive lung disease.^37–40^ Indeed, in a meta-analysis of nine studies totaling 4,699 participants in Bangladesh, Chile, India, Mexico, and Pakistan, higher arsenic exposure as measured by urine or drinking water concentrations was associated with reduced FEV_1_ and FVC, but not FEV_1_/FVC.^41^ In contrast, we found a relationship between arsenic and airflow obstruction (reduced FEV_1_/FVC), and statistically insignificant reductions in FEV_1_ and FVC. Relying upon spirometry to diagnose restrictive lung disease must be done with caution: total lung capacity (TLC) is the diagnostic gold standard to diagnose restrictive lung disease, and although FVC and TLC are related lung measurements, FVC-based algorithms unreliably identify restrictive lung disease.^42^

We did not find evidence that sweat chloride mediates the relationship between arsenic exposure and airflow obstruction, which suggests that there are mechanisms other than CFTR dysfunction that may explain arsenic’s associations with lung health. For example, arsenic has been shown to induce direct lung tissue injury in tracheal epithelial cells and alveolar macrophages through the generation of reactive oxygen species and stress proteins.^43,44^ Arsenic exposure is also associated with decreased serum CC16 levels,^45^ which is a lung protective protein secreted by alveolar epithelial cells that modulates inflammatory lung injury. Lastly, arsenic exposure is associated with increased matrix metalloproteinase-9, one of a class of proteins that regulate airway inflammation and have been implicated in the pathogenesis of obstructive lung disease.^46–48^

In a similar manner, better understanding of the mechanisms of arsenic toxicity may benefit individuals with genetically-determined cystic fibrosis. Not all the individuals in our study who were exposed to high concentrations of arsenic had elevated sweat chloride concentrations or lung dysfunction, suggesting that other factors may be associated with preserved epithelial chloride transport. These factors might reveal protective mechanisms that could be exploited to develop therapies for genetically-determined cystic fibrosis. The recent work in killfish and cell culture described above^20–22^ suggests such mechanisms might include suppressed cellular response of ubiquitinylation.

The strengths of our study include our direct measurements of individual-level long-term arsenic exposure using toenail clippings, sweat chloride concentrations, lung function, and – among those with elevated sweat chloride concentrations – genotyping to exclude pathogenic *CFTR* variants. Our study also has limitations. Firstly, as with all cross-sectional studies, because sweat chloride concentrations were measured at the same time that water and toenail samples were collected, we cannot confirm that arsenic exposure preceded their elevated sweat chloride concentrations. However, the prevalence of *CFTR* mutations in South Asian populations is low.^49^ This, in combination with the bench literature supporting CFTR dysfunction as a plausible mechanism through which arsenic exposure causes high sweat chloride concentrations, we believe our observations likely represent a true association with arsenic exposure. We also obtained PFTs in only a subset of the total study population because the COVID-19 pandemic interrupted PFT testing. However, there were no substantive differences in the participants who completed PFTs as compared to those who did not, so we do not anticipate that this biased our results. Finally, we were unable to characterize other pulmonary features of cystic fibrosis such as bronchiectasis, a finding that would be most evident on CT imaging as compared to plain films, or non-pulmonary features of cystic fibrosis such as pancreatic dysfunction. Future work directly measuring CFTR function in affected tissues including lung epithelium and neutrophils may be informative.

## CONCLUSIONS

Environmental arsenic exposure is associated with impaired CFTR function, as demonstrated by elevated sweat chloride concentrations, and airway obstruction. Potential applications of our study include future work aimed at development of a point-of-care sweat chloride test as a biomarker of arsenic exposure.

## Data Availability

All relevant data are within the manuscript and its Supporting Information files.

